# CRISPR-mediated functional mapping of *IL2RG* variants in primary human T cells predicts X-linked severe combined immunodeficiency

**DOI:** 10.64898/2026.04.27.26351884

**Authors:** Yvonne Rong, Mikhail Vysotskiy, Peixin Amy Chen, Ekansh Agrawal, Edward Marsh, Charlotte H. Wang, Daniel Carr, Rama Dajani, Brenna Gittins Maker, Furong Yu, Daniel B. Goodman, Eric Shifrut, Jennifer M. Puck, Alexander Marson, David N. Nguyen

## Abstract

Distinguishing pathogenic from benign mutation is critical for genetic diagnosis. A CRISPR-targeted saturation genome editing (SGE) platform in primary human cells assessed 489 single nucleotide variants (SNVs) in exon 5 of IL2RG, the gene causing X-linked SCID. The functional impact was clearly defined for 470 variants, agreeing with 100% (18/18) of ClinVar-deposited benign or likely benign annotations, and 100% (42/42) of pathogenic or likely pathogenic annotations. We discovered 90 novel loss-of-function mutations and validated an expected block in T-lymphocyte differentiation from edited hematopoietic stem cells.

---

Genetic diagnoses of inborn errors of immunity (IEI) are fraught with variants of uncertain significance (VUS) limiting efforts to distinguish a patient’s unique causative variant(s) from the abundant benign variants, particularly for missense mutations^1-3^. Severe combined immunodeficiency (SCID) is a rare, but prototypical IEI characterized by failure of hematopoietic stem cells (HSCs) to fully differentiate into mature T cells with subsequent lack of adaptive immunity ^4,5^. Untreated patients succumb to severe infections within the first years of life, with the standard of care being early allogeneic hematopoietic stem cell transplant (allo-HSCT). While SCID is now detected in many countries by newborn screening^6^, the genetic etiology is not always apparent. Even synonymous mutation has been shown to cause SCID^7^. Lack of genetic diagnosis is a barrier to family counseling, to excluding thymic or secondary T-cell deficiency that will not respond to allo-HSCT^5^, and to emerging attractive approaches of gene therapy or genome editing ^8-10^.

X-linked SCID (X-SCID) comprises approximately 30% of SCID cases ^11^ and is caused by loss-of-function (LOF) variants in the X-linked gene *IL2RG*, which encodes the common gamma chain (γc) of several cytokine receptors. *IL2RG* LOF prevents HSC differentiation into T cells and NK cells, and leads to intrinsic dysfunction in B cells ^12^. Most X-SCID cases are caused by single nucleotide variants (SNVs) in coding regions and splice sites, yet only 13% of all possible *IL2RG* coding variants have been annotated in ClinVar ^13,14^. When a patient presents with a variant not previously observed, incomplete knowledge of variant function could limit time-sensitive diagnostic certainty and delay patient care ^8^; without a molecular diagnosis gene and genome editing therapy would be precluded. These VUS dilemmas extend to the over >500 monogenic IEIs ^15,16^ and most other genetic disorders where patient variants are rare and unique (often private), underscoring the need for scalable forward genetics approaches to functional variant interpretation.

Experimental approaches to assess variant function *in vitro* (e.g., randomized oligo, PCR-based, or saturation mutagenesis) are historically limited by low throughput, disparate variant representation, narrow-spanning regions, and/or performance in physiologically and genetically dissimilar immortalized cell lines or yeast or mouse models ^17,18^. Base editing employed for high-throughput generation of mutations for functional analysis in primary human immune cells^19,20^ with wide screening capacity, but only a subset of possible mutations can be created by base editors. Computational pathogenicity prediction algorithms are reliant upon conservation scores (CADD, PolyPhen-2, SIFT) or utilize protein sequence and structural information (AlphaMissense, ESM1b). However, these tools often produce discordant predictions, lack fine-tuning for tissue- or cell-type-specific contexts, and are subjective to human interpretation biases limiting their clinical utility particularly for VUSs where interpretive demand is greatest^21-25^. Saturation genome editing (SGE) screens have emerged as a systematic experimental approach to interrogate the function of mutations in a gene or genomic region ^26,27^. Unlike other mutagenesis approaches, SGE introduces precise mutations into a user-specified genomic region in cells via CRISPR targeting; this allows measuring the individual mutation effect in a native genomic context with preserved regulation. Such screens have been valuable in mapping out the effects of variants previously unseen in patients and resolving VUS in *BRCA1, DDX3X, BAP1, CARD11*, and *RAD51C*. However, none have been performed in primary human immune cells^27-31^.

We hypothesized that impaired *IL2RG* function in primary human T cells would correlate with arrest of T-cell differentiation from HSPCs, although the exact molecular block to differentiation in X-SCID is yet to be fully defined^32^. Screening in T cells would also overcome numerical (eg coverage) limitations of comprehensive screening directly in HSPCs ^33^. Enabled by our recent advances in primary human cell engineering ^34-36^, we performed a CRISPR-targeted SGE screen of all possible SNVs in exon 5 of *IL2RG* directly in primary human T cells, using growth from IL-2 stimulus as a measure of functional effect. We focused on *IL2RG* exon 5 as it encodes part of the protein extracellular domain (Supplementary Figure 1), is the most frequently mutated in X-SCID patients^11,12^, and has the greatest number of unique annotated variants in ClinVar available for benchmarking screen results^14^.

## RESULTS

To assess the effects of all SNVs in exon 5 systematically, we generated a library of homology-directed repair (HDR) templates, each representing one of 489 different variants, that was electroporated together with targeting Cas9 ribonucleoproteins into T cells isolated from healthy (self-identified) male adult blood donors. As *IL2RG* is on the X chromosome, the use of male hemizygous donors enables precise editing of only one allele without compensatory effects from a second chromosome copy; each cell exhibits behavior due to only one variant. Edited T cells were expanded in culture with high doses of IL-2 to select for intact common gamma chain (γc) function (via IL-2 receptor) and sorted for γc surface expression (Fig. 1a). We collected T cells at 3 days and 14 days post-edit then sequenced genomic DNA (gDNA) and complementary DNA (cDNA). A functional score was defined as the log2 fold change between the relative frequency of each variant post-selection compared to the initial HDR template library. This score was adjusted similar to previously described methodology^27^ to account for sequencing depth, proximity to the nuclease cut sites, and averaged across two healthy cell donors (Supplementary Figure 2).

**Figure 1:**
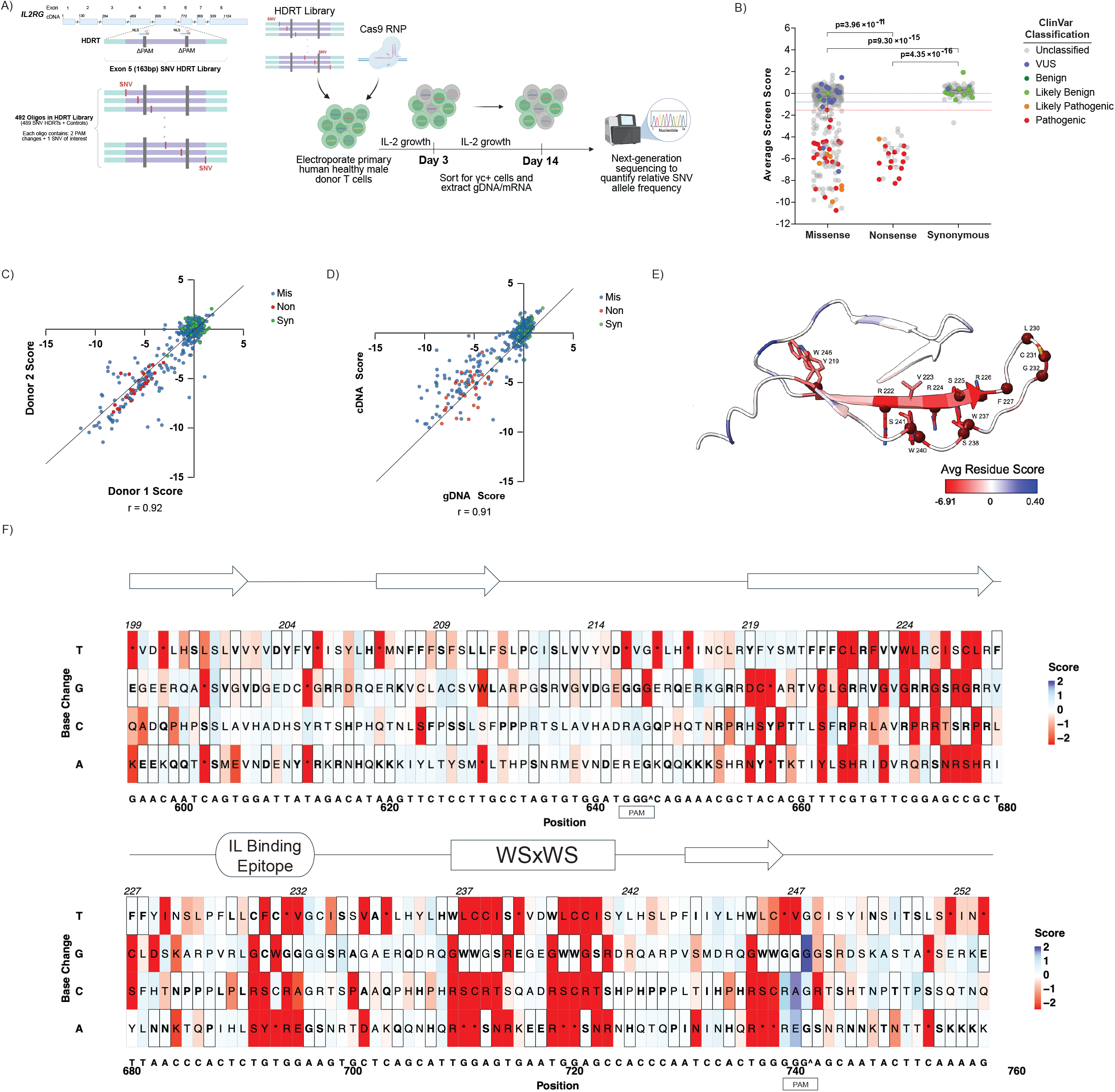
A saturation genome editing screen reveals impacts of IL2RG exon 5 variants on primary human T-cell fitness. **(A)** Schematic of CRISPR-targeted HDR insertion of SNV alleles enabling a comprehensive screen of IL2RG exon 5. A library of HDR templates, each containing one of 489 SNVs (plus two PAM-altering mutations) was electroporated along with two Cas9 RNPs into bulk (CD3+) T-cells from male donors. Cells were expanded under IL-2 stimulus, and at 3 days and 14 days post-edit, cells were flow-sorted for surface expression of the common gamma chain (γc). Bulk gDNA or mRNA was extracted for deep sequencing of SNV alleles, and the relative frequency of each variant post-selection determined a functional impact score based upon the log-fold change of the variant allele frequency compared to the initial HDRT library. **(B)** Functional scores of each SNV categorized by mutation type and further annotated with ClinVar (or HGMD) interpretation if available. Cutoffs for classification of variants into functional and non-functional categories depicted with variants scoring above the blue dashed line classified as “functional” (69.1%), variants below the red dashed line classified as “non-functional” (27.0%), and the rest are “intermediate” (3.9%). Scores are averaged across independent screens conducted in n=2 healthy cell donors. Error bars for each variant type represent the median and 25th/75th quantiles. P-value from Wilcoxon Test is shown. **(C-D)**: High correlation of functional scores between **(C)** average functional scores obtained from either cDNA or gDNA sequencing readouts displayed for each SNV labeled by mutation type, or **(D)**independent screen functional scores from gDNA sequencing readouts in n=2 healthy cell donors, displayed for each SNV and colored by functional score. **(E)** A functional score per amino acid for missense mutations is mapped onto the exon 5-encoded portion of γc (model generated by AlphaFold). Residues associated with known X-SCID mutations are highlighted with maroon spheres. **(F)** Heatmap of variant functional score by genomic position. Horizontal axis represents the genomic position in exon 5 of IL2RG with amino acid coordinates on top and cDNA coordinates on the bottom (numbered from the start codon). The GRCh38 assembly reference sequence (e.g. wildtype) is noted on top except for two synonymous mutations (marked with ^) introduced to alter the PAM sequence and block further editing. Vertical axis represents variant tested at each position. The resulting amino acid single letter code is shown in each box; bold letters with box outline are reference sequence. The color and intensity of the box represent the functional score for each variant with red indicating lesser fitness, and blue boxes representing greater fitness. A schematic of known structural regions in the portion of γc protein encoded by exon 5 are shown above the heatmap; arrows represent beta sheets.

**Figure 2:**
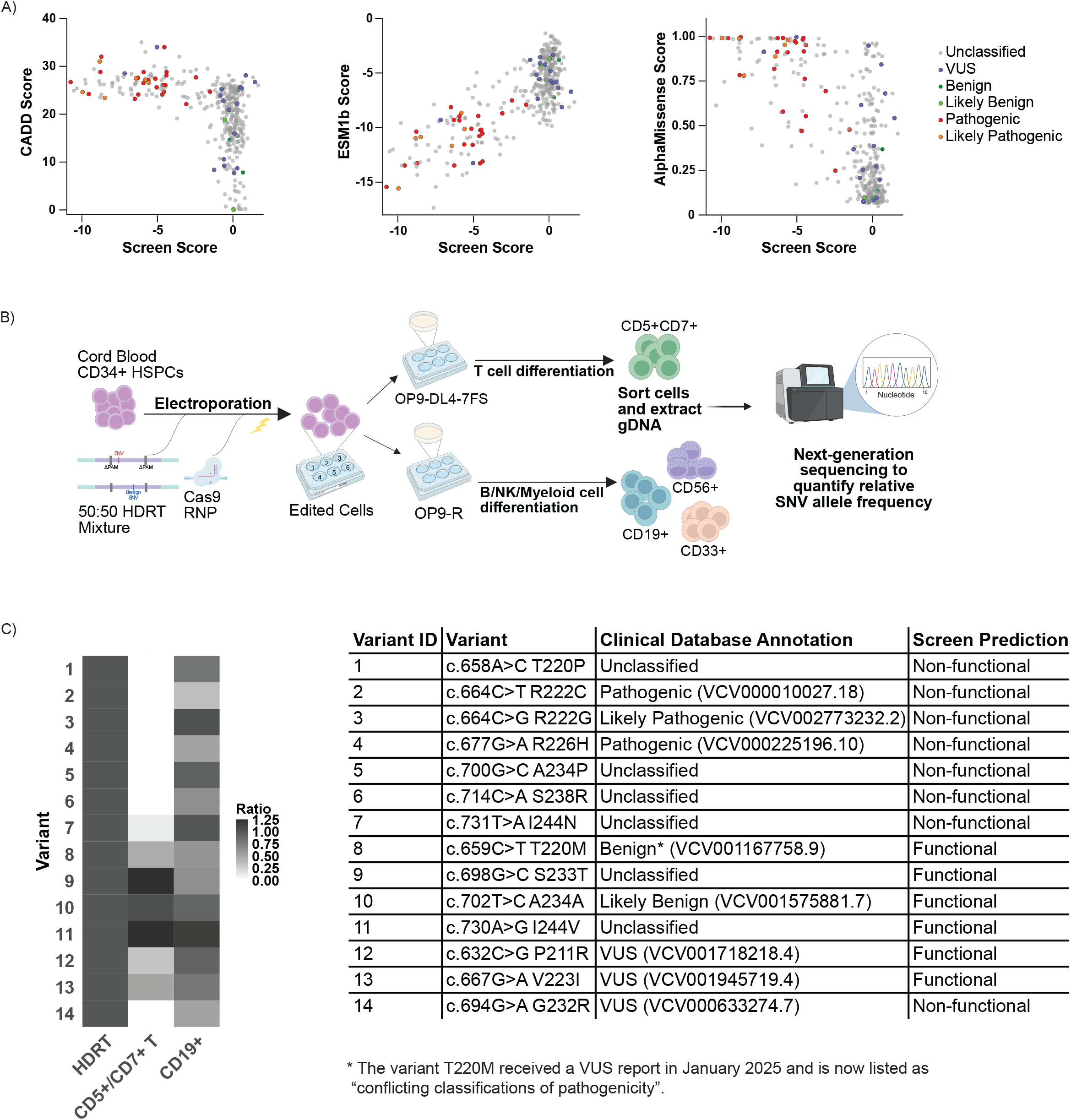
Comparison of SNV screen scores to variant effect prediction algorithms, and functional validation in HSPCs. **(A)** Comparison of each SNV screen functional scores to CADD score (left), ESM1B score (center), and AlphaMissense score (right), with label by ClinVar classification if available. Higher CADD and AlphaMissense scores and lower ESM1b score indicate greater likelihood of pathogenicity. AlphaMissense and ESM1b quantifies only missense variants. AlphaMissense scores are capped at 1. **(C)** Schematic of validation of variant impact upon HSPC differentiation into progenitor T or B cells. A 50:50 mixture of HDRTs recreating either the variant of interest or a ClinVar-benign variant (c.708T>C, H236H) was electroporated into cord blood CD34+ HSPCs from male donors, which were split for co-culture on the OP9-7FS or OP9-R lines that promote differentiation inv vitro into either T- or B-cell lineages, respectively. Cells were sorted for surface markers of maturation along each respective lineage, then gDNA was extracted and sequenced to calculate the relative frequency of either variant allele. The functional impact of a given variant upon HSPC differentiation into respective lineages is measured by the prevalence of alleles bearing the variant of interest relative to the benign variant. **(D)** Heatmap of adjusted ratios of alleles for fourteen selected test variants (relative to ClinVar benign variant, normalized to initial ratio in the HDRT mix) in sorted cell populations from HSPC differentiation: CD5+/CD7+ pre-T cells, and CD19+ B Cells. ClinVar accession numbers are listed where available.

We observed that nonsense mutations, as anticipated, had significantly lower functional scores than synonymous mutations (Fig. 1b), with the greatest separation at an early timepoint (3 days post-edit) sequenced using gDNA. The scores for this early-day screen are used for all the following analyses. The functional scores for T-cell growth were well-correlated between the two donors (Fig. 1d) and demonstrate a bimodal distribution (Supplementary Figure 3A). In addition, we compared scores from the early timepoint gDNA with cDNA sequencing (Fig. 1c), late timepoint (14 days) gDNA sequencing from the same biological donors, and a third donor that was not sorted for γc expression (Supplementary Figure 3). Across these readouts we observe similar nonsense-synonymous variant separation and similar distribution of missense variants. Using a mixture modeling approach to classify variants in the context of the bimodal distribution of the functional score ^27,37^, we separated variants into functional (69.1% of variants), nonfunctional (27.0% of variants), and intermediate (3.9% of variants) categories (Fig. 1B). All 29/29 nonsense variants fell into the nonfunctional category, and 94/95 synonymous variants fell into the functional category. The one nonfunctional synonymous variant restores a PAM site mutation that was introduced to prevent re-cutting by Cas9; this expected artifact leads to under-representation of this variant by decreasing the rate of variant knock-in. Of the 365 tested missense variants, 66.8% were classified functional, 27.9% nonfunctional, and 5.2% intermediate. An overview of functional scores by genomic position reveals certain regions more susceptible to loss of function when mutated (Fig. 1E,F); most of these regions correspond to protein structural regions with strong cross-species conservation (Supplementary Figure 4). For example, our unbiased screen clearly identifies the WSxWS motif that is highly conserved across metazoans and is found in many cytokine receptors with a critical role in function and signal activation ^38,39^.

We tested whether our T-cell functional screen correctly classified *IL2RG* mutations previously found in patients that have been annotated as X-SCID-causing (pathogenic) in the ClinVar or the Human Gene Mutation Database (HGMD). All 5/5 *IL2RG* variants annotated as “benign” in ClinVar were classified in the functional category (Fig. 1b). We accurately classify 13/13 “likely benign” and 7/7 “likely pathogenic” *IL2RG* variants. 35/35 *IL2RG* mutations annotated as “pathogenic” (in either database) are classified as nonfunctional in our SGE approach. Of the 18 variants annotated as VUS in ClinVar, many of which have some corroborating clinical evidence, we predict that 15 are functional and 2 are nonfunctional, with the remaining classified as “intermediate” (Supplemental Table 1).

We compared the scores and nominal mutation classifications in *IL2RG* exon 5 from our *in vitro* functional screen to computational tools commonly used for predicting variant pathogenicity ^24,40-43^ (Fig. 2a). The correlation in scores was high between the SGE screen and CADD v1.7 (absolute Pearson 0.6, Spearman’s rho 0.63), Alpha Missense (0.68, 0.57), and ESM1b (0.78, 0.63); correlation with Polyphen (0.45, 0.39) and SIFT (0.32, 0.45) were low (Supplemental Figure 5). We further compared these tools to our SGE dataset regarding the ability to classify database-annotated variants within *IL2RG* exon 5. There are dozens of missense variants predicted as nonfunctional by CADD and AlphaMissense that we classify as functionally benign in T cells. One ClinVar-benign *IL2RG* variant had a CADD score above 20, placing it in the top 1% of predicted deleterious variants (c.659C>T, T220M), which was correctly classified as T-cell functional using SGE screen data. While AlphaMissense misclassified one annotated pathogenic variant (R226H c.677G>A) as benign, one annotated benign variant (Q200H c.600A>C) as ambiguous, and two annotated pathogenic variants (R224W c.670C>T, R222C c.664C>T) as ambiguous, our SGE screen provided the correct classifications for each; one variant, c.689T>C L230P, was intermediate in both the screen and AlphaMissense. Notably, 14% of *IL2RG* exon 5 variants evaluated by AlphaMissense were labeled as “ambiguous”, as opposed to 4% (n= 19/489) in our screen data. Overall, our SGE screen data improves classification quality over existing computational methods, demonstrating a benefit of direct empirical evidence over variant effect prediction.

We next verified that screen-classified nonfunctional or functional mutations affect primary human T-cell growth. We individually assessed 28 different *IL2RG* exon 5 variants in hemizygous T cells introduced by CRISPR-targeted knock-in with a 1:1 mixture of two HDR templates, one containing the specific variant of interest and the other containing a ClinVar annotated benign synonymous mutation (H236H c.708T>C) as a functional control. We followed the relative frequency of the two variants over 8 days of culture with or without IL-2 to compare the fitness of each variant. All ClinVar or HGMD annotated pathogenic variants nearly disappeared by day 3 (Supplementary Figure 6), and all but one screen-identified nonfunctional variant (W246S, c.737G>C) showed similar dropout. Either annotated benign or screen-predicted functional variants persisted in T cells through day 8 at levels similar to the H236H control.

We further assessed if the functional screens of *IL2RG* variants in T cells would be predictive of function in primary human HSPCs, where LOF mutations block differentiation into T cells while permitting B-cell development: the (T-B+) phenotypic hallmark associated with X-SCID (Fig. 2b) ^12,44^. Attempts to perform the entire exon 5 screen directly in HSPCs were limited by poor resolution of functional and non-functional mutations, with multiple factors likely contributing to poor library coverage (Supplemental Discussion and Supplemental Figures 7-9). We performed a focused arrayed validation of screen-identified variants similar to the competition assay validation studies performed in T cells. We selected variants falling into five categories for validation studies: those previously unannotated and predicted nonfunctional by our screen (A234P c.700G>C, S238R c.714C>A, T220P c.658A>C, I244N c.731T>A), previously unannotated and predicted functional by our screen (S233T c.698G>C, I244V c.730A>G), and controls with annotated X-SCID pathogenic or likely pathogenic mutations (R226H c.677G>A, R222C c.664C>T, R222G c.664C>G) or annotated benign mutations (A234A c.702A>C, T220M c.659C>T). We also selected several ClinVar-annotated VUS to test: G232R c.694G>A (screen-predicted non-functional), V223I c.667G>A (screen-predicted functional), and P211R c.632C>G (screen-predicted functional). To control for stochastic failure of individual edited HSPC clones to differentiate, we similarly introduced *IL2RG* exon 5 variants into primary human CD34+ HSPCs by CRISPR-targeted knock-in with a 1:1 mixture of a specific variant of interest and the ClinVar benign synonymous mutation (H236H c.708T>C). Edited HSPCs were then differentiated *in vitro* by co-culture with the OP9 and OP9-DL4-7FS line ^45^ (Fig. 2b). Compared to H236H, all annotated and screen-predicted nonfunctional mutations were depleted in CD5+/CD7+ pre-T cells by week 3 of T-cell differentiation while cells bearing annotated or screen-predicted functional variants differentiated into pre-T cells at similar rates (Fig. 2c). Edited HSPCs bearing any of the variants, including annotated pathogenic or screen-predicted nonfunctional mutations, were able to differentiate into CD19+ B cells on the OP9 line ^46,47^. We note that the variant T220M was later classified in ClinVar as a VUS after another data deposit; however, unimpaired T cell differentiation from HSPCs bearing this T220M mutation is consistent with its initial classification as benign (ClinVar Variation ID: 1167758 Accession: VCV001167758.9). Four variants (G232R, A234P, A234A and S233T were differentiated to the CD4+/CD8+ T-cell stage via co-culture on OP9-7FS cell line, and also into CD56+ NK cell progenitors and CD33+ myeloid progenitors via co-culture on the OP9 cell line. Consistent with expectations, the annotated and screen-predicted pathogenic variants were highly depleted in NK cells, while all four variants remained present in myeloid cells (Supplementary Figure 10,11,12). While G232R and A234P persisted at reduced frequency at the CD5+/CD7+ stage, they were fully-depleted by the CD4+/CD8+ stage.

Interestingly, R222C c.664C>T, an annotated X-SCID mutation that has been associated with atypical “leaky” SCID phenotype (Tlow/− B+ NK+) ^13^, was largely depleted in the CD5+/7+ cell population, comparable to annotated pathogenic and screen-predicted nonfunctional variants (e.g., R226H). Taken together, our validation experiments show that novel T-cell screen-predicted *IL2RG* exon 5 LOF variants indeed block T-cell development *in vitro*, and across multiple different mutations we observe that this effect occurs at or before the pre-T (CD5+/7+) stage of development.

In summary, we applied SGE to systematically and comprehensively map the functional impact of all 489 possible SNVs in exon 5 of *IL2RG* in the native genomic context in primary human T cells. Using a primary human T-cell fitness readout, we accurately classified 97% of database-annotated X-SCID mutations imparting decreased T-cell fitness, 100% of annotated benign variants with no significant effect on T-cell fitness, and, without bias, identified critical structural elements and known high sequence conservation. We further validated selected SNVs in primary human HSPCs for blockade of T cell differentiation that could be indictive of X-SCID pathogenicity. We report experimental predictions for the function of all SNVs, including nonfunctional mutations that may be identified in future X-SCID patients. Our approach is also successful in potentially reclassifying VUSs, an otherwise slow process requiring the accumulation of patients or direct experimental investigation conducted with patient samples ^2,3^. Our SGE screen approach, first in T cells and then validated in HSPCs, directly identifies both non-functional and functional variants allowing it to be used as an experimental assay for formal variant classification in the PS3/BS3 evidence categories ^48^. One apparent strength of directly assaying *IL2RG* mutations in primary cells for either normal or impaired function is avoiding autonomously growing cancer-derived cell lines that often bypass autoregulation of the JAK-STAT pathway ^49, 50^. This SGE approach generates a high-resolution, accurate functional data set directly in primary human cells that will improve timely genetic diagnoses for patients with X-SCID. In parallel with ongoing advances in computational mutation effect prediction, datasets such as our SGE screen can provide empirical training and benchmarking data, especially for diseases where clinically validated genes and variants are limited. We anticipate that SGE screening in primary cells will a generalizable approach for predicting IEI variant function and to pinpoint mutations likely to benefit from expanding gene therapy and genome editing opportunities.

## METHODS

### Guide RNA selection

Candidate guide RNAs (gRNAs) targeting *IL2RG* exon 5 were selected from CRISPOR based on predicted efficiency, off-target effects, and the ability to create synonymous change(s) to the PAM/protospacer codons. Through an initial empirical testing, multiple gRNA pairs were assayed for CRISPR-targeted knock-in editing using a homology-directed repair template (HDRT) with multiple synonymous mutations spanning the exon and validated by gDNA sequencing. The top gRNA pair (TCTCCTTGCCTAGTGTGGAT, GGAGCCACCCAATCCACTGG) were selected and used for all further SGE screen and validation work performed in this paper.

### Cas9 ribonucleoprotein (RNP) production

For initial gRNA testing and SGE screens, CRISPR RNA (crRNA) and trans-activating crRNA (tracrRNA) were synthesized (IDT Technologies), resuspended in IDT duplex buffer at a concentration of 160 µM, and stored in aliquots at -80°C until use. For validation experiments, sgRNAs were synthesized (IDT Technologies), resuspended in IDT duplex buffer at a concentration of 80 µM, and stored in aliquoted at -80°C until use. To make gRNA, crRNA and tracrRNA aliquots were thawed, mixed at a 1:1 volume ratio, and annealed by incubating at 37°C for 30 min to form an 80 µM gRNA solution; sgRNA were thawed and used immediately.

Poly-L-glutamic acid (PGA) (Sigma-Aldrich) was mixed with the gRNA solution (crRNA/tracrRNA or sgRNA) at a 0.8:1 volume ratio, followed by complexing with Cas9-NLS (40 µM, Berkeley Macrolab) at a 1:1.8 volume ratio for a final molar ratio of 2:1 gRNA to Cas9-NLS. The gRNA/PGA/Cas9-NLS complex was incubated at 37°C for 15 min to form a 14.3 µM RNP solution. RNP solution was aliquoted and stored at -80°C until use. At the time of electroporation, RNP was thawed and incubated at 37°C for 5 min before use.

### HDR Template Library Design and Production

The *IL2RG* Exon 5 HDRT library was designed with two PAM altering mutations (c.645G>A, c.741G>A) incorporated into each template (at the optimal gRNA cut sites) in the library to prevent re-cutting of the Cas9 nuclease and to use as a marker for downstream data analysis.

A custom script then created a list of variant alleles altering one base position to each of the three other nucleotides across the entire exon. The custom library was synthesized as gBlocks (IDT Technologies) and cloned by NEBuilder assembly (New England Biolabs) into a custom pUC19 vector with pre-defined homology arms of length ∼350 bases flanking each side of *IL2RG* Exon 5 for targeting homology-directed repair. The plasmid library was sequence verified by NGS, and analyzed as described below in “SGE Screen Data Analysis” to ensure equal distribution of each SNV. A linear dsDNA HDRT library was then amplified via PCR using KAPA HiFi polymerase (Kapa Biosystems) with the addition of truncated Cas9 targeting sequences (tCTS) at both ends as previously described ^34^, purified using SPRI beads (1X), and resuspended in water to 0.5–2 μg/μl measured by light absorbance on a NanoDrop spectrophotometer (Thermo Fisher Scientific) for subsequent knock-in via electroporation.

For validation studies in T cells and HSPCs by electroporating a 1:1 mixture of a test variant and an annotated benign synonymous variant (H236H), the same two PAM altering mutations and 250-base homology arms used in the *IL2RG* Exon 5 HDRT library were incorporated into each individual HDR template. The individual HDR templates were synthesized as gene fragments (Twist Bioscience) and PCR amplified and purified as described above for the HDRT library.

### Isolation and culture of primary human T cells

Human peripheral blood leukopaks enriched for PBMCs (STEMCELL Technologies) were collected from healthy self-identified male donors. Primary human CD8+ T cells were isolated from fresh PBMCs using the EasySep Human CD8+ T cell Isolation Kit STEMCELL Technologies) following the manufacturer’s protocol. Primary human CD3+ T cells were isolated from fresh PBMCs using the EasySep Human T cell Isolation Kit (STEMCELL Technologies) following the manufacturer’s protocol.

For 48 hours prior to electroporation, freshly-isolated T cells were activated with anti-human CD3/CD28 Dynabeads (Thermo Fisher Scientific) at a bead-to-cell ratio of 1:1 at a starting density of 1 million cells/mL, and cultured in X-VIVO 15 (Lonza) supplemented with 5% fetal bovine serum, 50 µM 2-mercaptoethanol, IL-2 (300 U/mL), IL-7 (5 ng/mL), and IL-15 (5 ng/mL) (PeproTech).

### Primary T cell electroporation

CD8+ T cells were used for the SGE screen, while CD3+ T cells were used for validation of selected SNVs. Activated primary T cells were collected from their culture vessels, diluted in MACS buffer (1X volume), resuspended to break apart cell-bead clusters, and transferred to 50 mL conical tube(s). To separate T cells from anti-human CD3/anti-CD28 Dynabeads (Thermo Fisher Scientific), cells were placed in an EasySep cell separation magnet (STEMCELL Technologies) for 5 min and de-beaded cells were collected in new 50 mL conical tube(s).

Immediately before electroporation, de-beaded cells were centrifuged at 100xg for 8-10 min and resuspended in P3 Primary Cell Nucleofector Solution with Supplement 1 (Lonza). Each experimental condition received 500,000 activated de-beaded T cells resuspended in 20 µL of P3 buffer solution, 3.5 µL of RNP (50 pmol), and 2 µL of HDR template (0.5 µg total) unless otherwise stated. For the SGE screen in T cells, an HDRT library spanning *IL2RG* exon 5 (described above) was used. For the validation studies in T cells, a 1:1 v/v mixture of the test variant and annotated benign synonymous variant (H236H) HDRTs was used. All T cell electroporation experiments were carried out in 96-well format using a Lonza 4D-Nucleofector 96-well system with pulse code EH-115.

Immediately after electroporation, 80 µL of pre-warmed X-VIVO 15 (no cytokines) was added to each electroporation well and incubated at 37°C for 10-15 min. For the SGE screen, all electroporated T cells were transferred to a T-25 flask and diluted in fresh X-VIVO 15 supplemented with IL-2 (500U/mL) to achieve a nominal concentration of 1 million cells/mL. Every 2-3 days, T cells were counted and split to achieve a nominal concentration of 1 million cells/mL in fresh X-VIVO 15 supplemented with IL-2 (500 U/mL) in new T-25 flask(s). On days 3 and 14 post-electroporation, ¼ of cells were stained for flow cytometry analysis, ½ of cells were used for gDNA and cDNA sequencing, and the remaining cells were added to fresh X-VIVO 15 supplemented with IL-2 (500 U/mL) in new T-25 flask(s). For validation studies, 500,000 cells were electroporated and split into triplicates of 166,666 cells in 200uL (0.83×10^6 cells/mL) of X-VIVO 15 media supplemented with IL-2 (500U/mL). Every 2-3 days, T cells were counted and maintained at 0.83×10^6 cells/mL in new 96-well plates. On day 3 post-electroporation, one replica plate was used for flow cytometry and gDNA sequencing. For the remaining two plates, one replica plate was maintained in X-VIVO 15 supplemented with IL-2 (100U/mL) and the other replica plate was maintained in X-VIVO 15 without IL-2 for further studies. On days 5 and 8 post electroporation, for both replica plates with and without IL-2 supplementation, a portion of cells were stained for flow cytometry analysis, and a portion of cells were used for gDNA sequencing. Refer to sections “Flow Cytometry and Cell Sorting” and “Preparation of gDNA and cDNA for Next-Generation Sequencing” for more details.

### HSPC culture

Human cord blood CD34+ cells were collected from healthy self-identified male donors (STEMCELL Technologies). HSPCs were thawed in a 37°C water bath, transferred to a conical tube, and 10 mL of warm IMDM was added dropwise to the cells while gently vortexing. Cells were spun at 200xg for 10 min, resuspended in 500 μL of warm IMDM supplemented with 0.1mg/mL DNase I, and incubated for 15 min at RT. The cells were then rescued with 9 mL of warm IMDM, counted, centrifuged at 200xg for 10 min, and resuspended in 1 mL warm StemSpan SFEM-II (STEMCELL Technologies) supplemented with Penicillin 100 U/mL Streptomycin 100 μg/mL and additional cytokines: stem cell factor (SCF) at 100 ng/mL, thrombopoietin (TPO) at 100 ng/mL, and Flt3 at 100 ng/mL (PeproTech). Cells were plated in TC-treated 6-well plates (Thermo Fisher Scientific) at a concentration of 0.25×10^6 cells/well at a total volume of 2 mL and incubated at 37°C for 48 hours prior to electroporation. Cord blood aliquots from multiple donors were pooled to achieve sufficient starting numbers of cells.

### HSPC electroporation

For each condition in the validation studies, a 1:1 v/v mixture of the test variant and the ClinVar benign synonymous reference (H236H) HDR templates was prepared at 0.25 µg/µL. Immediately before electroporation, approximately 48 hours post-thaw, HSPCs were transferred to 50 mL conical tube(s), centrifuged for 10 min at 100xg, and resuspended in electroporation buffer P3 (Lonza). Each experimental condition received 250,000 HSPCs resuspended in 18 µL of P3 buffer, 4 µL of RNP (58 pmol), and 2 µL of HDR template (0.5 µg total). All HSPC experiments were carried out in 96-well format using a Lonza 4D-Nucleofector 96-well system with pulse code ER-100. Immediately after electroporation, cells were rescued by adding 80 µL of warm StemSpan SFEM-II media (no cytokines) into each well and incubated at 37°C for 10 min. Cells were transferred to a 12-well TC-treated plate, diluted to a concentration of 250,000 cells/mL in StemSpan SFEM-II media supplemented with stem cell factor (SCF) at 100 ng/mL, thrombopoietin (TPO) at 100 ng/mL, and Flt3 at 100 ng/mL (Peprotech), and cultured at 37°C for 24 hours. A 10,000-cell aliquot of each condition was preserved for pre-differentiation sequencing, and the remaining cells were then equally divided for co-culture on OP9-DL4-7FS or OP9-R mouse stromal cells. The HDR template mixtures were preserved for sequencing.

### Differentiation of edited HSPCs by OP9-DL4-7FS and OP9-R co-culture

Edited (or control) HSPCs were differentiated as previously described ^45,51^ either into T cell progenitors on the OP9-DL4-7FS, or into B cell, NK cell, and myeloid progenitors on the OP9-R cell line; both cell lines were obtained as a gift from the laboratory of Juan Carlos Zúñiga-Pflücker. OP9 media was prepared by supplementing α-MEM (Minimum Essential Medium Eagle Alpha Modification) with 5% (v/v) FBS and Penicillin 100 U/mL–Streptomycin 100 μg/mL (ThermoFisher Scientific) and sterile-filtered for the maintenance of OP9-DL4-7FS and OP9-R cells. OP9-DL4-7FS co-culture media is the same as OP9 media. OP9-R co-culture media was prepared by supplementing α-MEM with up to 20% (v/v) FBS and Penicillin 100 U/mL– Streptomycin 100 μg/mL (ThermoFisher Scientific) and sterile-filtered.

Approximately 1-2 weeks prior to co-culture with edited (or control) HSPCs), OP9-DL4-7FS and OP9-R frozen vials were thawed in a 37°C water bath, transferred to a 50 mL conical tube(s), and 10 mL warm OP9 media was added dropwise to the cells while gently vortexing. Cells were centrifuged at 500xg for 5 min, resuspended in 1 mL warm OP9 media, counted, and transferred to 6-well TC-treated plate(s) at 1.5×10^5 cells per well in 3 mL fresh OP9 media total. Every 3-4 days, cells were washed with 1X PBS (Cytiva), treated with 1 mL 0.05% Trypsin-EDTA (Thermo Fisher Scientific), counted, centrifuged at 500xg for 5 min, and replated at 3-5 ×10^4 cells per well in 2-3 mL fresh OP9 media in 6-well TC-treated plates. Passaging schedule and cell seeding density were adjusted accordingly as to not exceed 90% cell confluency.

One day before co-culture with edited (or control) HSPCs, either OP9-DL4-7FS or OP9-R plates were washed with 1X PBS (Cytiva), treated with 1 mL 0.05% Trypsin-EDTA (Thermo Fisher Scientific), counted, centrifuged at 500xg for 5 min, and seeded at 1.5×10^5 cells per well in 3 mL of respective co-culture media in 6-well TC-treated plates to achieve ∼90% confluency in each well. The next day edited (or control) HSPCs were then added directly onto OP9-DL4-7FS or OP9-R monolayers at 5×10^4 cells/well for each condition. For co-culture with OP9-DL4-7FS, no additional cytokines were added. For co-culture with OP9-R, IL-7 (5 ng/μL), Flt3 (5 ng/μL), and SCF (30 ng/μL) were added to the culture media. The co-cultures were incubated for 7 days at 37°C and passaged every 7 days.

Differentiating HSPCs were isolated from the monolayers every 7 days, then re-plated onto fresh OP9-DL4-7FS or OP9-R monolayers that had been seeded and cultured for 24 hours as described above. For the first 2 weeks of differentiation, the co-culture cells (both HSPCs and OP9-DL4-7FS or OP9-R cells) were treated with 1 mL 0.05% Trypsin-EDTA (Thermo Fisher Scientific) and filtered through a 40-μm nylon mesh into a 50 mL conical tube. After week 2 of differentiation, 0.05% Trypsin-EDTA (Thermo Fisher Scientific) was no longer used, and co-culture cells were disaggregated by thoroughly pipetting medium and filtered through a 40-μm mesh into a 50 mL conical tube. Cells were then centrifuged at 550xg for 5 min, resuspended in respective co-culture media, counted, and seeded onto fresh OP9-DL4-7FS or OP9-R monolayer at 0.5×10^6 cells/well in a 6-well TC-treated plate in 3 mL total of respective co-culture media. For co-culture with OP9-R, IL-7 (5 ng/μL), Flt3 (5 ng/μL), and SCF (30 ng/μL) was supplemented to the co-culture media. The co-cultures were incubated at 37°C.

### Flow cytometry and cell sorting

Flow cytometric analyses were performed on an Attune NxT Flow Cytometer with a 96-well autosampler (Thermo Fisher Scientific). Flow sorting was performed on a FACSAria (BD Biosciences). Cell surface staining for flow cytometry was performed by centrifuging at 400xg for 5 min, resuspending cells in 30 µL of FACS buffer (1-2% FBS and 1µM EDTA in PBS) with antibodies diluted accordingly, and incubating for 30 min at RT in the dark. After incubation, cells were washed once with FACS buffer before resuspension and analysis. For SGE screen T cell analysis and sorting, cells were stained with anti-human Fc blocking solutions, live-dead stain and surface-marker targeting antibodies (anti-human CD4-Brilliant Violet 421, CD8-APC, CD132-PE) (Supplemental Figure 8).

For analysis of HSPC differentiation, cells differentiating into progenitor T cells on the OP9-DL4-7FS were stained with anti-human and anti-mouse Fc blocking solutions, live-dead stain, anti-human CD45-Brilliant Violet 421, CD34-PE-Cy7, CD7-APC, and CD5-PE between weeks 1-3 of differentiation; with anti-human and anti-mouse Fc blocking solutions, anti-human CD45-Brilliant Violet 421, CD7-APC, and CD5-PE for sorting at 3 weeks of CD5+CD7+ pro-T cells; with anti-human and anti-mouse Fc blocking solutions, live-dead stain, anti-human CD45-Brilliant Violet 421, CD3-PE, CD4-APC, and CD8-PE-Cy7 at weeks 4-6 of differentiation; and with anti-human and anti-mouse Fc blocking solutions, anti-human CD45-Brilliant Violet 421, CD3-PE, CD4-APC, and CD8-PE-Cy7 for final sorting of CD4+CD8+ pre-T cells (Supplementary Figures 10,11).

Cells differentiating into progenitor B cells, NK cells, and myeloid cells on the OP9-R were stained with anti-human and anti-mouse Fc blocking solutions, live-dead stain, anti-human CD45-Brilliant Violet 421, CD34-PE-Cy7, CD33-PE, CD19-APC, and CD10-Brilliant Violet 711 between 1-3 weeks of differentiation; and with anti-human and anti-mouse Fc blocking solutions, anti-human CD45-Brilliant Violet 421, CD33-PE, CD19-APC, and CD56-PE-Cy7 for final sorting (Supplementary Figures 10,11). All cytometry data were processed and analyzed on FlowJo software (BD Biosciences).

### Preparation of gDNA and cDNA for next-generation sequencing

After the saturation genome editing in T cells and differentiation studies in HSPCs, sorted cells were pelleted, and either genomic DNA was extracted via phenol-chloroform isopropanol precipitation or mRNA was stabilized in TRIzol (Thermo Fisher Scientific). For gDNA extraction, cells were lysed overnight in buffer (1% SDS, 50mM Tris PH 8.0, 10mM EDTA, and NaCl) and then treated with 10mg/mL RNAse A (Qiagen) and 20 mg/mL Proteinase K (Thermo Fisher Scientific). Genomic DNA was purified from the cell lysate using phenol-chloroform isopropanol precipitation. For cDNA preparation, cells were lysed and mRNA was purified using a Direct-zol RNA Miniprep Plus kit (Zymo Research) according to the manufacturer’s instructions and subsequently converted to cDNA using the Omniscript RT kit (Qiagen) or Maxima H Minus First Strand cDNA Synthesis kit with oligo(dT) (Thermo Fisher Scientific) according to the manufacturer’s instructions.

A three-step PCR approach was used to amplify sequences from either gDNA or cDNA, and then add adapters and indexes for next-generation sequencing. For gDNA, the first PCR (PCR#1) amplified a 1443bp region of *IL2RG* using PCR primers placed well outside of the edge of the HDRT homology arms, using KAPA HiFi HotStart polymerase for 15 cycles, followed by a 1.0X SPRI bead purification. The PCR primers are AAAAAGAAAAGCAAAGTGGACCT and AGGGACGTGAATGTAGCAGC. For cDNA, PCR#1 was not required.

A second PCR (PCR#2) amplifies *IL2RG* exon 5 and attaches (Nextera) P5 and P7 Illumina sequencing adaptors using KAPA HiFi HotStart polymerase for 15-20 cycles, followed by a 1.4X SPRI bead purification. The PCR#2 primers (without adapter sequences) are TTGGGCTCATGGATTGGGTC and AGTATGTTTTAATTCTCCCTTCTCTCA for gDNA and TGAACCACTGTTTGGAGCACT and CCAACAGAGATAACCACGGC for cDNA. The third PCR (PCR#3) attaches unique i5 and i7 indices using NEB Next Ultra II Q5 polymerase for 15 cycles, followed by a 1.0X SPRI bead purification. All PCR amplicon concentrations were quantified via Quant-iT PicoGreen dsDNA Assay (Thermo Fisher Scientific) or Qubit.

Normalized samples were pooled and sequenced on an Illumina NextSeq 1000/2000 with a 2×150 bp reads run mode at the UCSF CAT Core or Innovative Genomics Institute NGS Core.

### SGE screen data analysis

For each sample, paired reads were merged using FLASH2 with an extended maximum overlap (-M 165). Cutadapt was used to trim reads to the exon only, and CRISPResso2 was used to enumerate mutations in each sample and map to a library of inserted templates {https://doi.org/10.14806/ej.17.1.200} ^52^ (Supplementary Figure 2).

For the SGE screen, edgeR ^53^ was used to calculate log_2_ fold changes (LFC) between variant counts in each sample and variant counts in the original HDR template library. Data sets were processed and adjusted independently for gDNA or cDNA sequencing.

To adjust for potential bias around the Cas9 cut sites, a LOESS (locally estimated scatterplot smoothing) model was fit to the LFCs within the range of synonymous variants with span=0.15, and the predicted values were subtracted from the log_2_ fold change for each SNV according to position. The position-adjusted scores were linearly scaled such that the median synonymous and median nonsense SNVs for each donor would match the median synonymous and median nonsense SNV values averaged across donors. The donor average of these normalized values for each SNV was used as a functional score. To classify variants into the functional / nonfunctional / intermediate categories (where non-functional is lower fitness / depletion), a two-component Gaussian mixture model (mixtools package in R) was fit on scores for synonymous and nonsense variants ^37^. From the fitted model parameters, a posterior probability function was defined to compute, for any given score, the probability that it belonged to the non-functional component. The “non-functional” component was defined to have score values lower than the functional population. The non-functional population retains a posterior probability of P (non-func) ≥0.99, while the functional population has a P (non-func) ≤ 0.01; all variants of 0.01 > P (non-func) < 0.99 are classified as “intermediate”. This analysis methodology was adapted from Findlay et. al ^27^. The two synonymous variants which directly restored the PAM changes (c.656A>G G215G and c.741A>G G247G) were excluded from the loess and mixture modeling steps due to bias in knock-in and potential artifactual dropout of the synonymous variants. Analysis code is available at https://github.com/nguyenlab-igi/SGE Conservation scores for each residue in exon 5 were obtained from ConSurf and mapped onto the AlphaFold model of *IL2RG* using ChimeraX ^54-56^.

### Validation of SNV effect data analysis

For validation experiments in T-cells and HSPCs, competing a variant of interest against a clinically validated benign SNV, raw gDNA sequencing reads were processed as above using Flash2, Cutadapt, and CRISPResso2 (Supplementary Figure 13). The effect of each variant was quantified by the ratio of the test variant count to the c.746T>C p.H236H (ClinVar benign variant), normalized by the ratio of the variants in the starting HDRT mixture. T-cell validation studies were performed on two donors; the average ratio is reported unless one donor had missing data.

### Clinical databases and prediction algorithms

ClinVar classifications are current to March 2025 ^14^. HGMD data and various predictive algorithm scores were accessed in October 2024 ^40,57^. cDNA position numbering is from the first position in the *IL2RG* reference mRNA sequence.

## Supporting information

Supplementary Figures and Discussion

## Data Availability

All data produced in the present study are available upon reasonable request to the authors

## Acknowledgements

OP9-7FS, OP9-DL1, and OP9-R cell lines were a gift from Dr. Juan Carlos Zúñiga-Pflücker of the University of Toronto.

DNN was supported by NIH grants L40AI140341, K08AI153767, and P01AI138962. JMP received funding from the California Institute for Regenerative Medicine CLIN-17127; UCSF Living Therapeutics Initiative LTI-7032103; UC Berkeley, UCLA, and UCSF Danaher Beacon initiative MCA-12046; NIH grant P01AI138962. The Marson laboratory has received research support from the Parker Institute for Cancer Immunotherapy, the Emerson Collective, Arc Institute, Juno Therapeutics, Epinomics, Sanofi, GlaxoSmithKline, Gilead and Anthem and reagents from 10x, Ultima, Genscript, Illumina, Cella, and NIH grant P01AI138962.

Flow sorting was performed at the UCSF Parnassus Flow Cytometry Core (RRID:*SCR_018206*) supported in part by Grant NIH P30 DK063720 and by NIH 1S10OD021822-01. Sequencing was performed at the UCSF CAT, supported by UCSF PBBR, RRP IMIA, and NIH 1S10OD028511-01 grants.

