## Supplementary Figures and Discussion for "CRISPR-mediated functional mapping of *IL2RG* variants in primary human T cells predicts X-linked severe combined immunodeficiency"

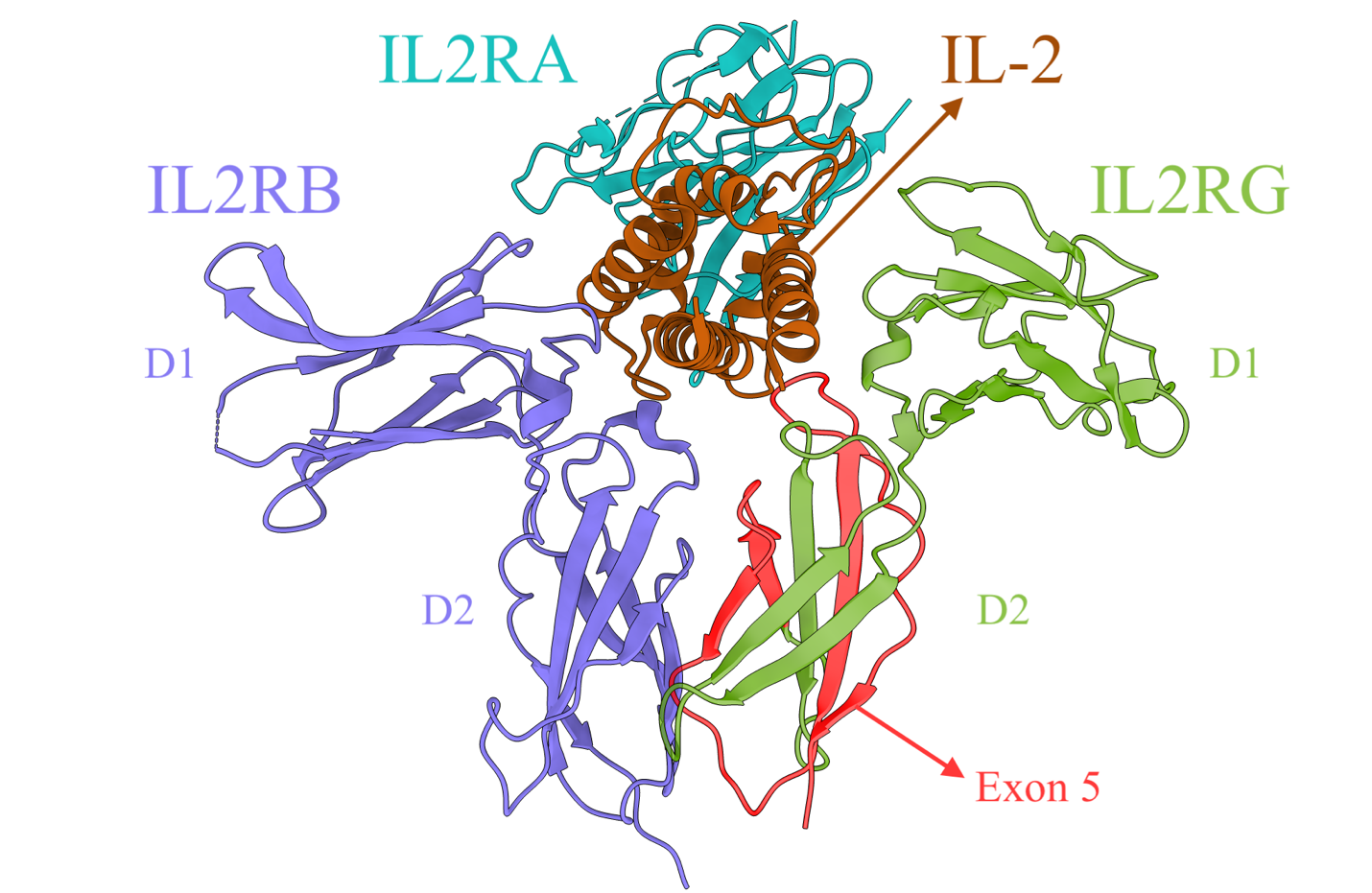

Supplementary Figure 1. Location of exon 5 within protein product of *IL2RG*. IL-2 receptor common gamma chain (IL2RG) (green) is shown in complex with the IL-2 receptor alpha chain (IL2RA) *(teal)* , IL-2 recetpor beta chain (IL2RB) (blue*),* and with bound IL-2 (brown) (PDB: 2ERJ). Portion encoded by *IL2RG* Exon 5 is highlighted in red. D1 and D2 refer to fibronectin domains (PMID: 16293754). Structure annotated in Chimera.

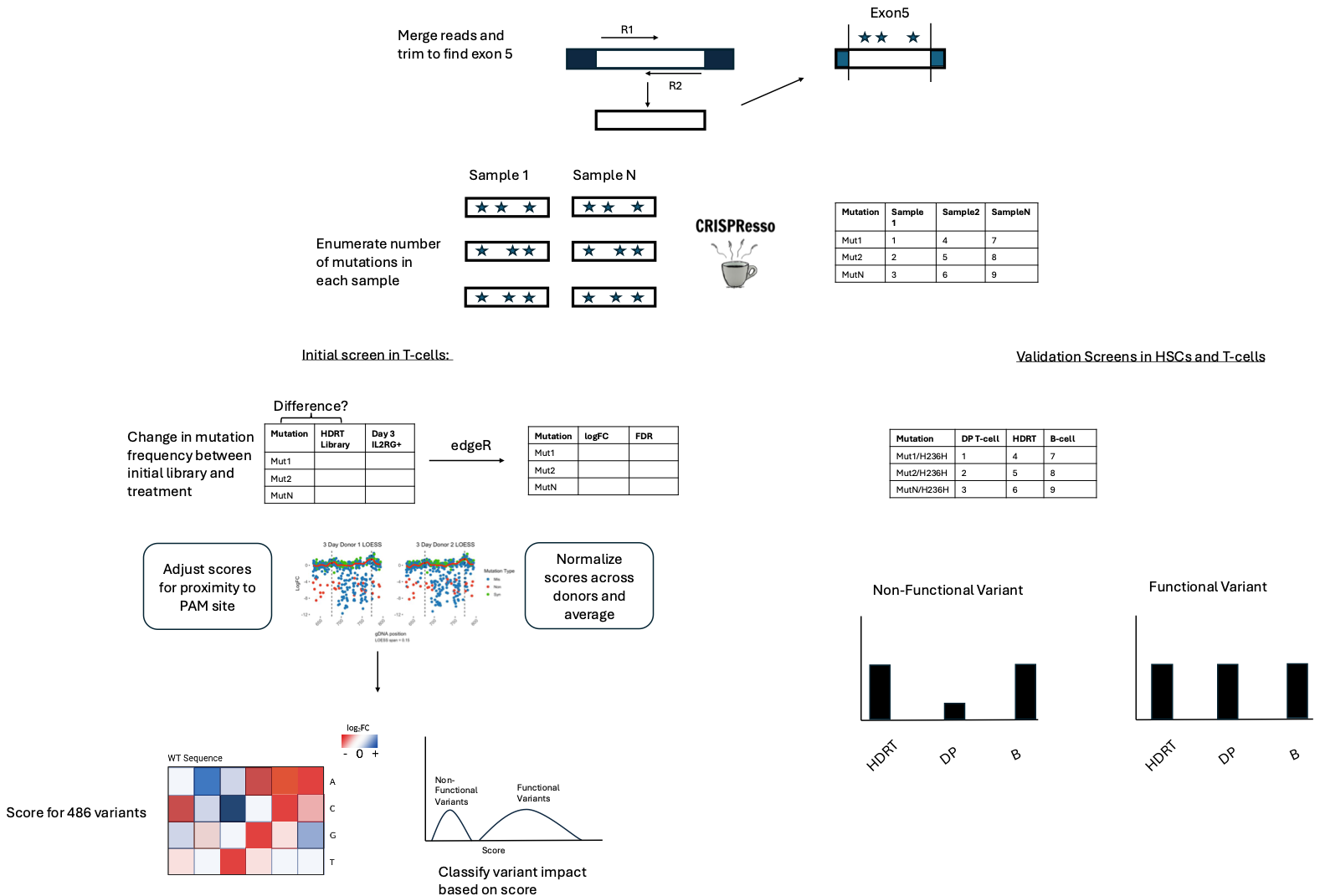

Supplementary Figure 2A: A graphical overview of the computational pipeline established for pre-processing sequencing reads to trim down to only the sequence representing exon 5, then alignment and allele counting facilitated by CRISPresso2, initial calculations of fold change in relative SNV allele frequency between screen outcome and initial HDR template library, post-processing adjustment and normalization to achieve a fitness score, and classification of variant function.

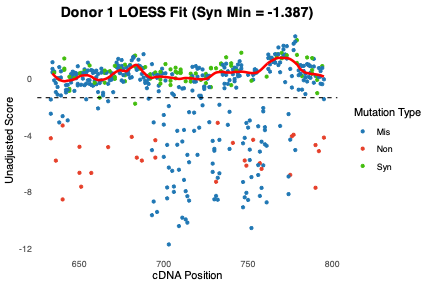

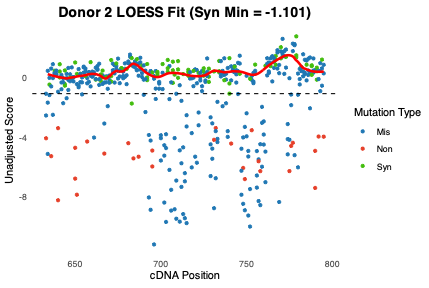

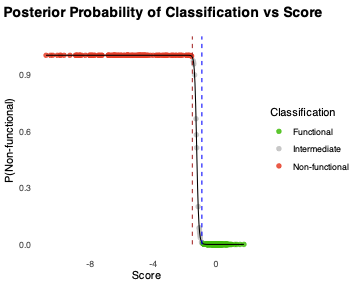

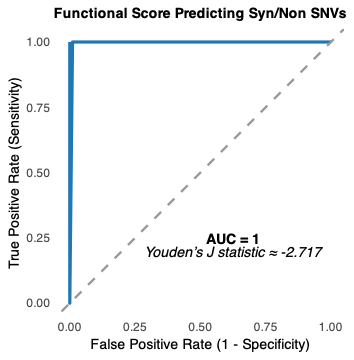

Supplementary Figure 2B: **(Top)** Log fold change (between baseline HDRT library and cells sorted at day 3) of sequenced allele counts per variant plotted by genomic position. A LOESS-smoothed curve (red line) enables calculating a functional score with correction for the positive peak in LFC occurring close to the Cas9 RNP cut sites, which is an artifact of locally greater propensity for SNV knock-in HDR proximal to the double strand break. **(Middle)** A two-component Gaussian mixture model was fit to a dataset filtered for nonsense and synonymous variants. For each variant, the posterior probability of being non-functional was computed from the relative contribution of the non-functional Gaussian density to the total density from both mixture components at that score. Variants to the left of the red dashed line (function score = -1.47) were categorized as ‘non-functional’, variants to the right of the blue dashed line (function score = -0.83) were categorized as ‘functional’, and the remaining variants were classified as ‘intermediate’. The red and blue cutoff lines correspond to scores where P(non-functional) = 0.99 and P(non-functional) = 0.01, respectively. **(Bottom)** The Receiver Operating Characteristic (ROC) curve shows perfect discrimination of nonsense and synonymous mutations by functional score, with 100% sensitivity and specificity at score -2.717 (calculated by maximizing Youden’s J statistic).

(A)

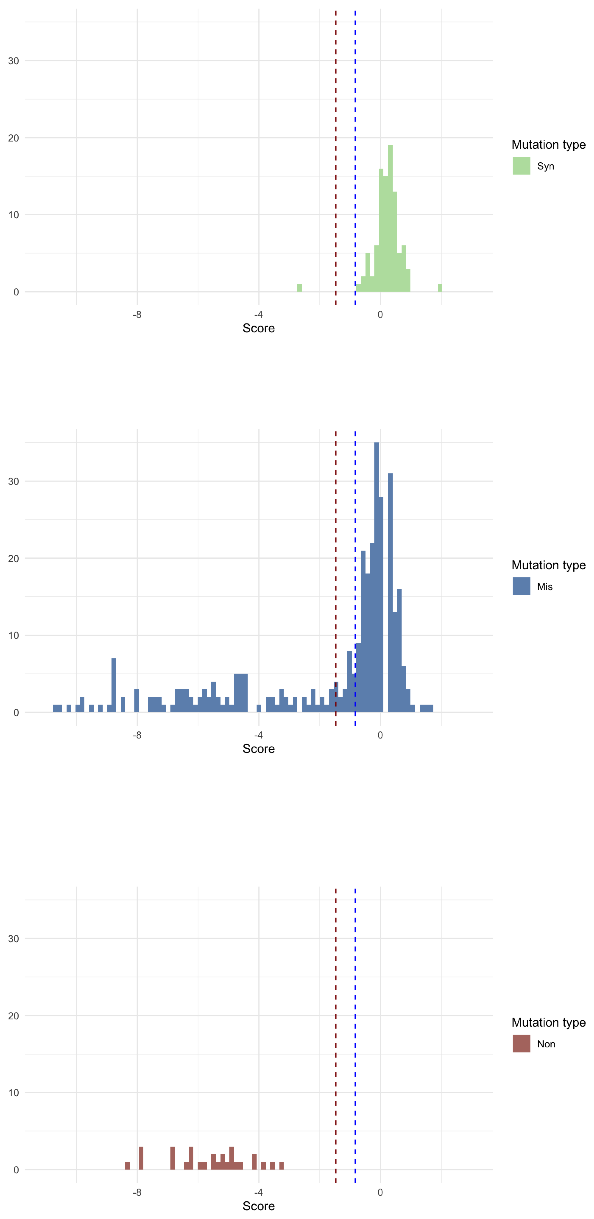

(B)
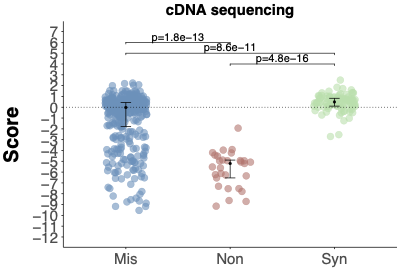

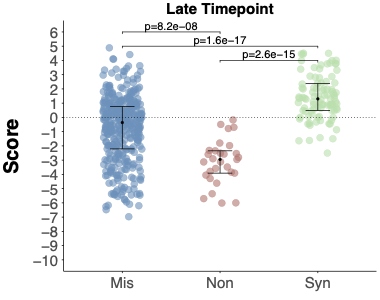

(C)

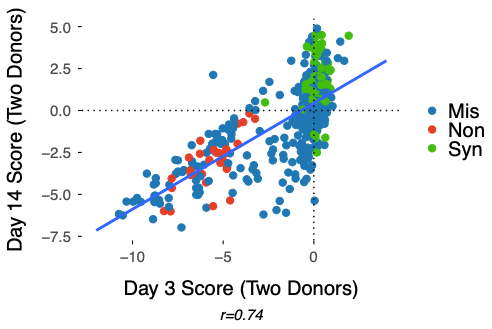

(D)
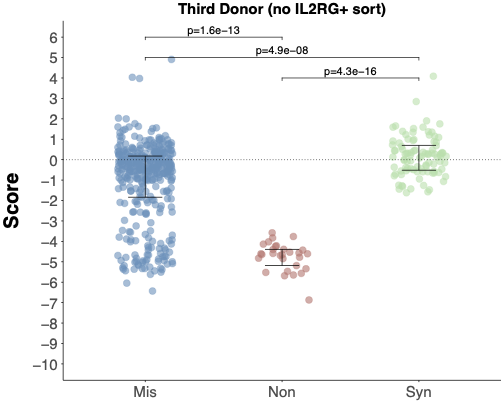

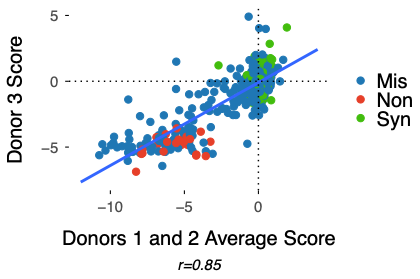

Figure S3: (A) Histogram representation of screen scores for each SNV averaged across n=2 donors separated by mutation type. (B) Distribution of screen scores for each SNV averaged across n=2 donors per mutation type using data derived from cDNA sequencing at the 3 day timepoint (left) or gDNA sequencing at the 14 day time point (right). (C) Comparison of scores between the early and late time points. (D) (Left) Distribution of screen scores per mutation type using data derived from gDNA sequencing of a third donor with screen performed with IL-2 growth only, without γc+ sorting. Cutoffs for classification of variants into functional and non-functional categories depicted with variants scoring above the blue dashed line classified as “functional”, variants below the red dashed line classified as “non-functional”, and the variants between classified as “intermediate”. Error bars represented median and quartiles, p-values are from Wilcoxon Test. (Right) Comparison of scores between the two donors reported in the main screen and the third (unsorted) donor.

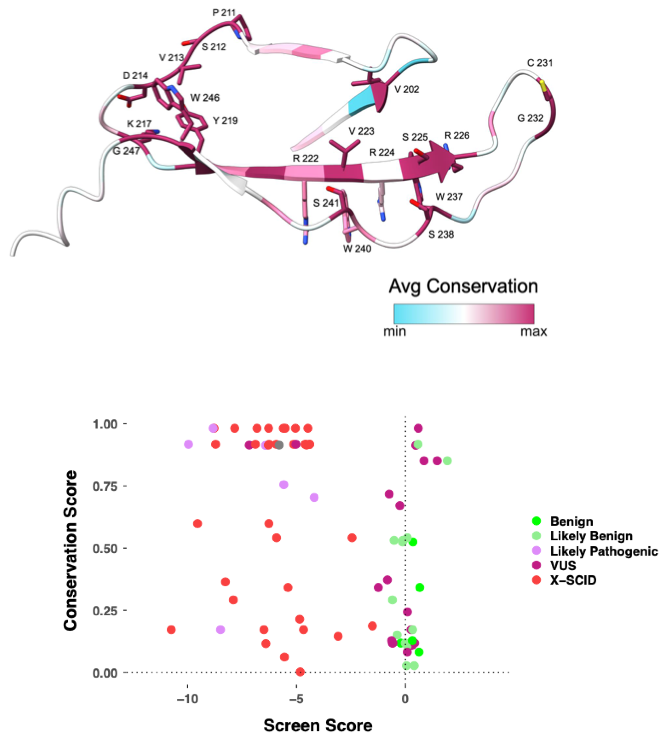

Figure S4: Screen results compared to conservation scores in *IL2RG*.

Top left: Amino acid conservation scores per amino acid mapped onto predicted portion of the yc structure encoded by Exon 5. Side chains of the most conserved residues are displayed. Model generated from Consurf and AlphaFold 2,3.

Top right: Functional Scores for all missense mutations averaged per amino acid mapped onto predicted portion of the yc structure encoded by Exon 5. Residues with the lowest Functional Score are displayed. Residues associated with X-SCID mutations Clinvar are highlighted with maroon spheres. Model generated from AlphaFold. Bottom: Scatterplot of the conservation score for variants annotated in clinical databases compared to screen functional score averaged per amino acid.

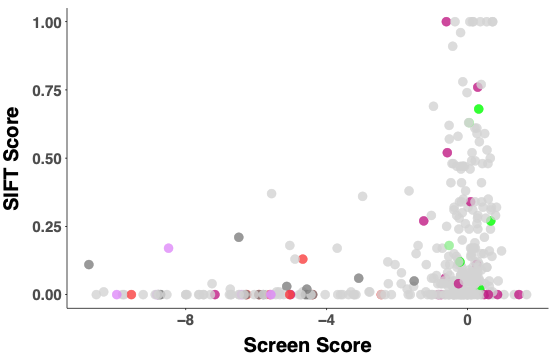

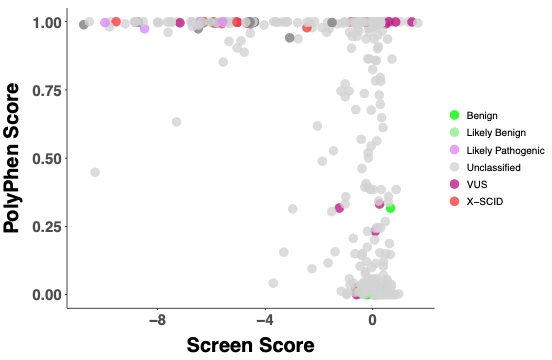

Figure S5: Comparison of each SNV screen functional score to SIFT score (left) and PolyPhen score (right), with label by with ClinVar classification if available. PolyPhen score closer to 1 and SIFT score closer to 0 indicates higher likelihood of pathogenicity. Only missense variants are represented.

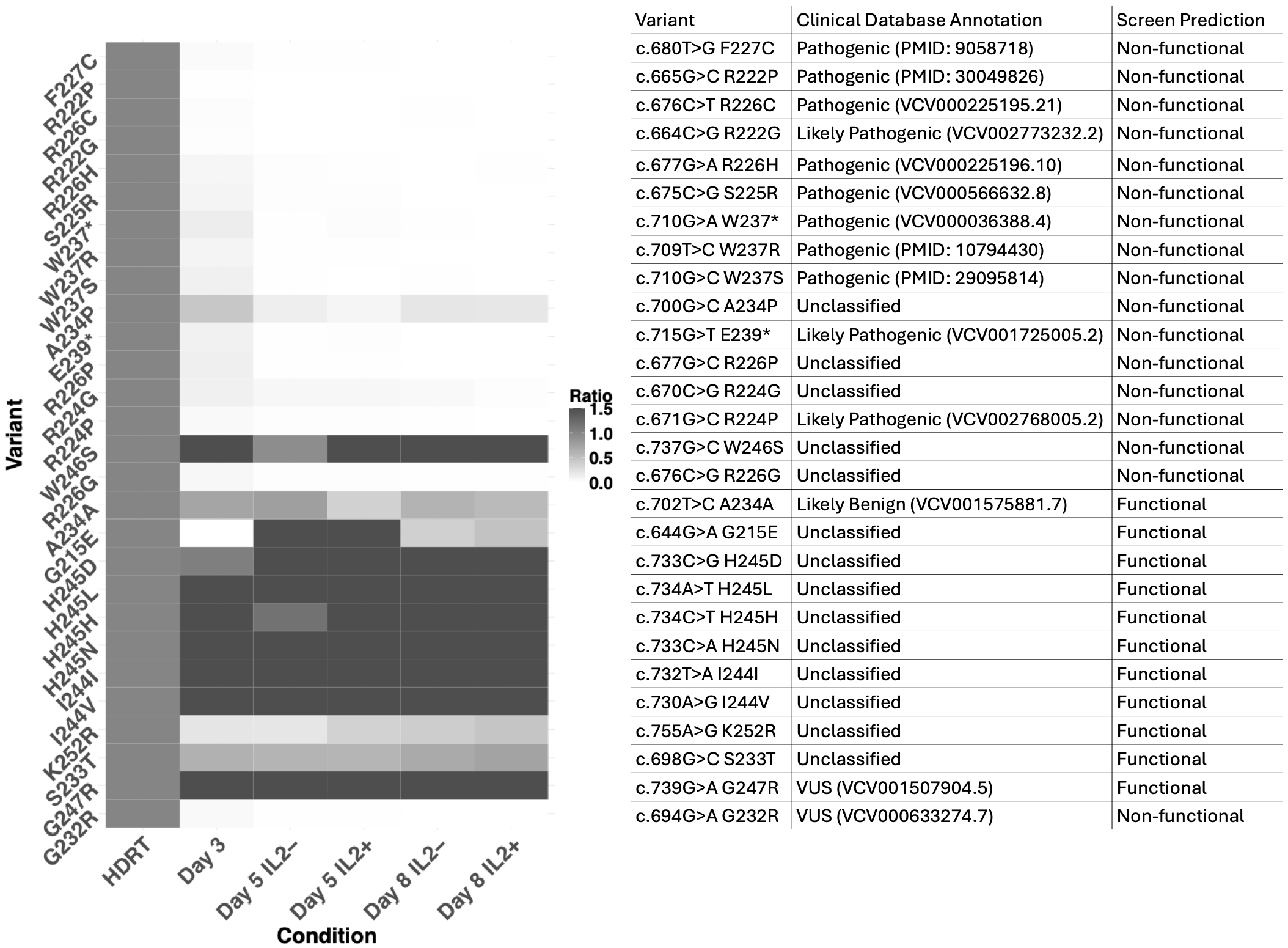

Figure S6: Screen predictions of 28 test variants were validated by competing a variant of interest with a known benign variant through T cell growth with and without IL2 supplementation. A 50:50 mixture of HDRTs recreating the variant of interest and a ClinVar-benign variant (c.708T>C, H236H) was electroporated into primary human CD3+ (bulk) T cells and grown for 3 days in the presence of IL-2, split, and then further cultured with or without IL-2 supplementation. At indicated time points, gDNA was extracted and sequenced to calculate the relative frequency of either variant allele. The functional impact of a given variant upon T cell growth is measured by the prevalence of alleles bearing the variant of interest relative to the benign variant. (Left) Heatmap of adjusted ratios of alleles for selected test variants (relative to ClinVar benign variant, normalized to initial ratio in the HDR template) (Right) List of 28 test variants. ClinVar accession numbers or reference publication PubMed IDs are provided if available.

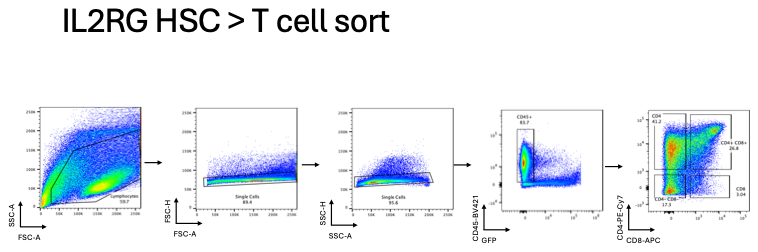

Figure S7.

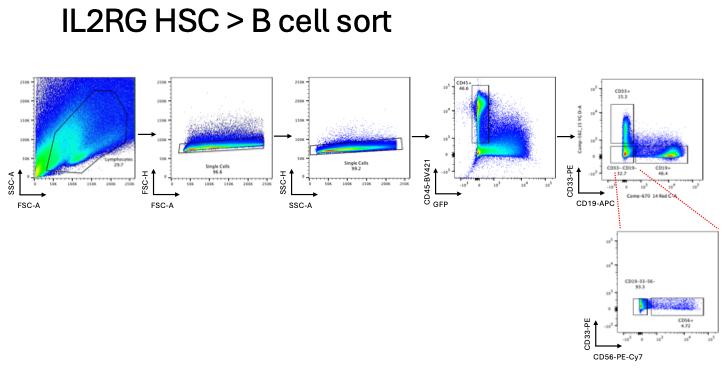

Figure S8.

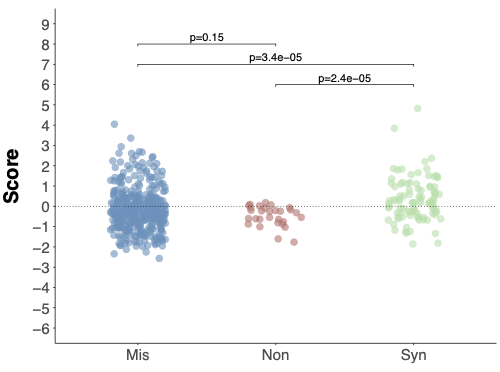

Figure S9

Figures S7-9: The full library of HDR templates, each containing one of 489 SNVs along IL2RG exon 5 (plus two PAM-altering mutations), and two Cas9 RNPs were

electroporated into cord blood CD34+ HSPCs, which were split for co-culture on the OP9-DL-4 or OP9-R lines that promote differentiation inv vitro into either T- or B-cell lineages, respectively (SFig 7-8). Edited HSPCs differentiated for 6 weeks on OP9-DL-4 were sorted for CD4+ CD8+ pre-T cell populations with gating strategy as shown (SFig 7). Edited HSPCs differentiated for 3 weeks on OP9-R were sorted for CD19+ pre-B cell, CD33+ myeloid lineage, or CD56+ NK-cell progenitor populations with gating strategy as shown (SFig 8)..

Genomic DNA was extracted for deep sequencing of SNV alleles, and the relative frequency of each variant post-selection determined a functional impact score based upon the log fold change of the variant allele frequency compared to the initial HDRT library. Variants were scored using the same methodology as T-cell screens. (SFig 9) Functional scores of each SNV categorized by mutation type. Error bars represented median and quartiles. P-value is given from Wilcoxon test.

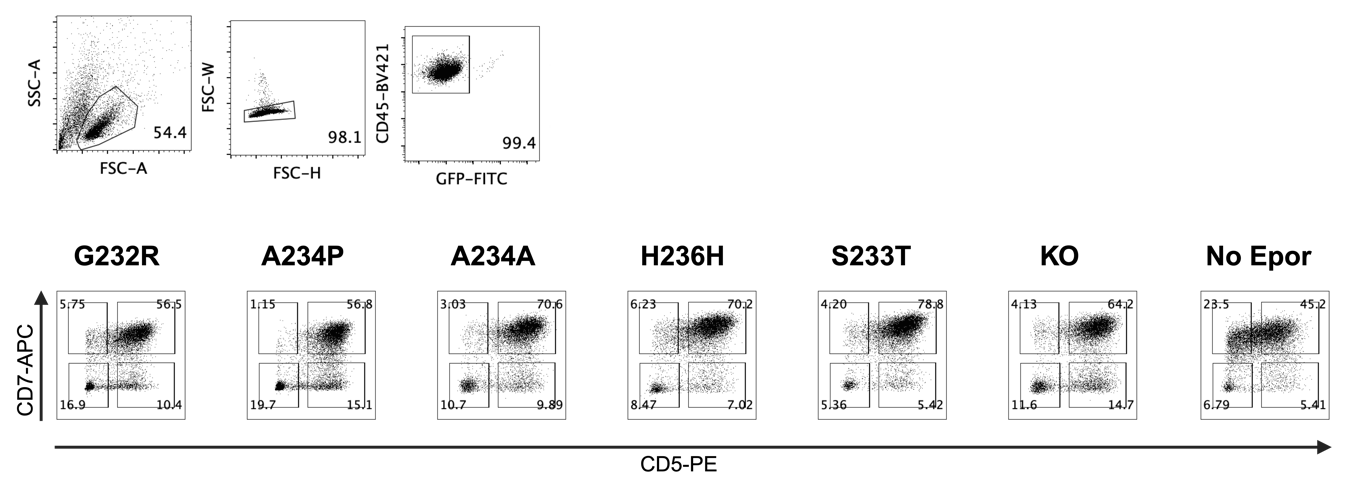

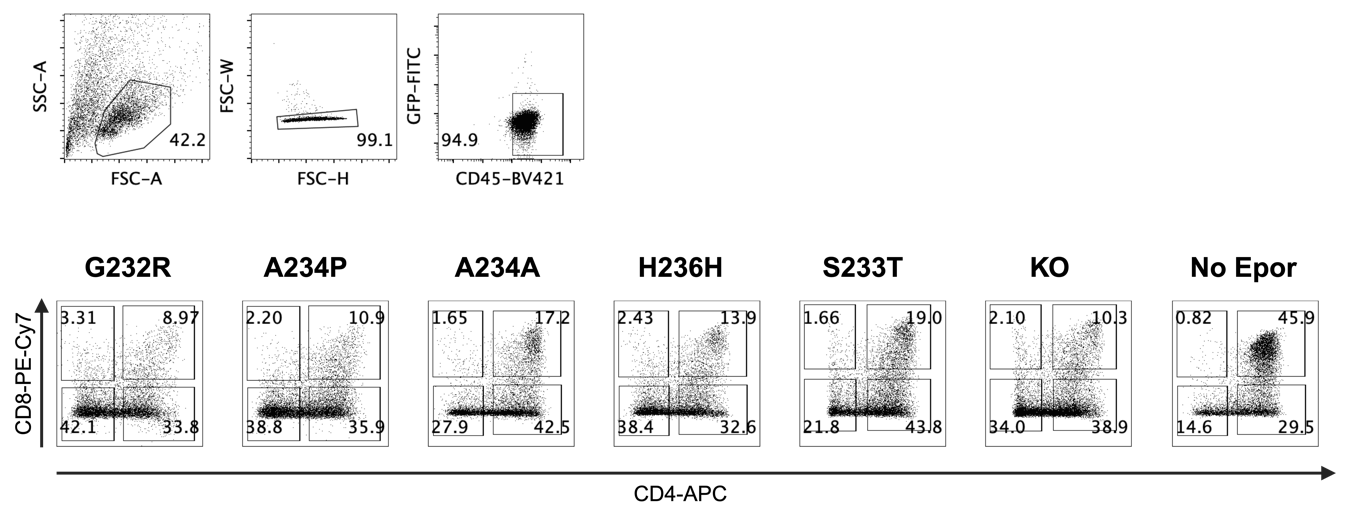

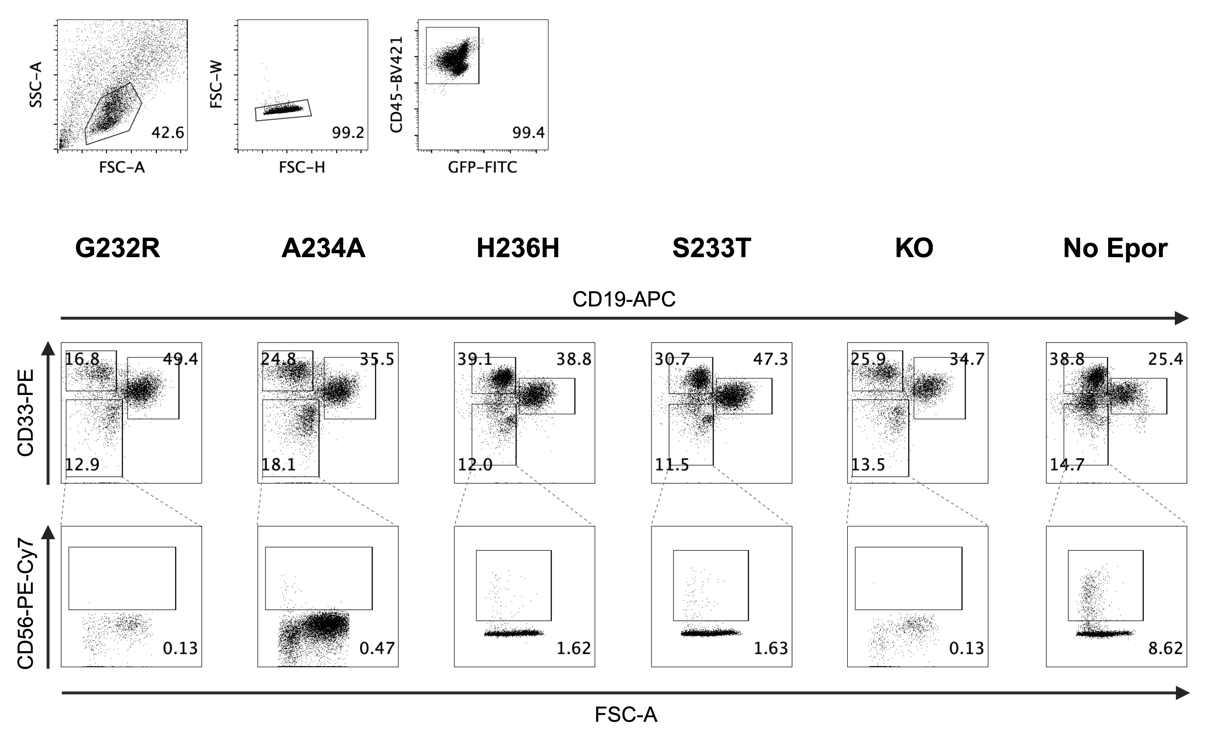

Figure S10: *Representative flow cytometry gating strategy and sort gates from Week 3 T-cell lineage differentiation (top), Week 6 T-cell lineage differentiation (middle), and Week 3 B/myeloid/NK-lineage differentiation (bottom) during HSPC validation studies of selected variants on interest or controls (KO = knockout, No Epor = treated similarly (“mock”) but without electroporation.*

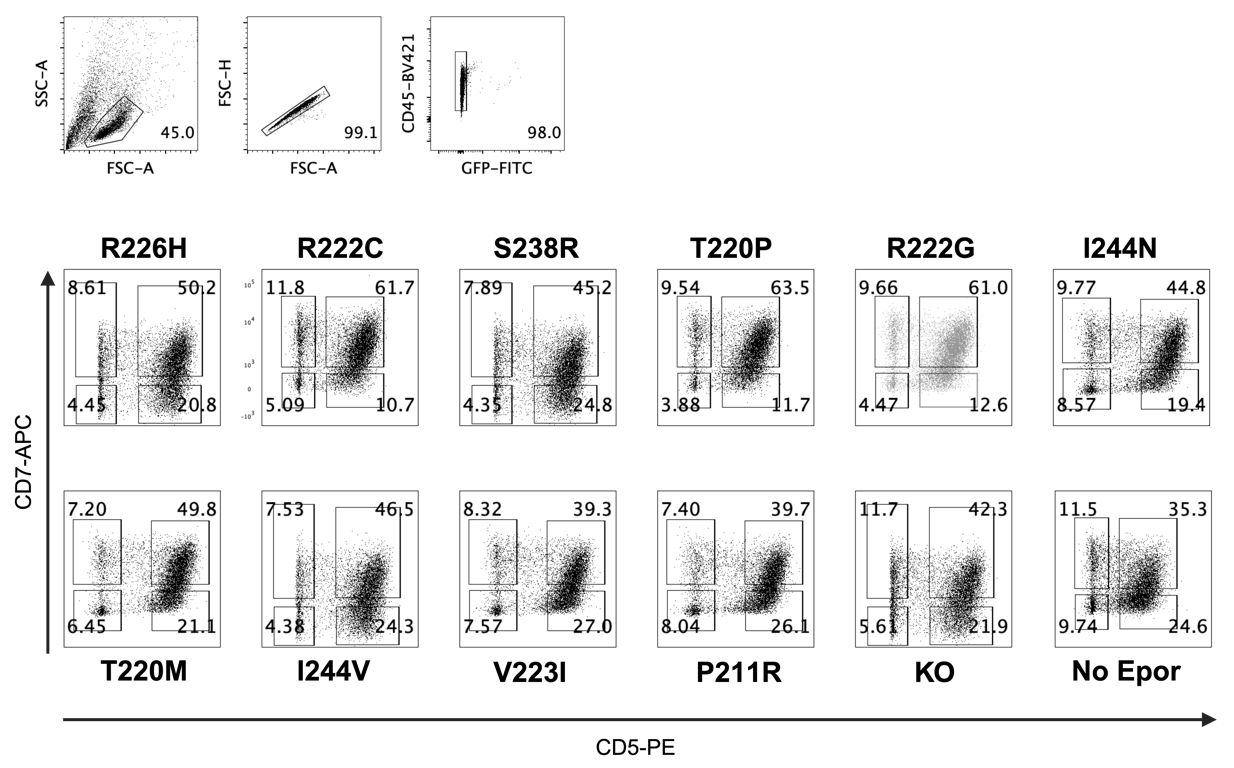

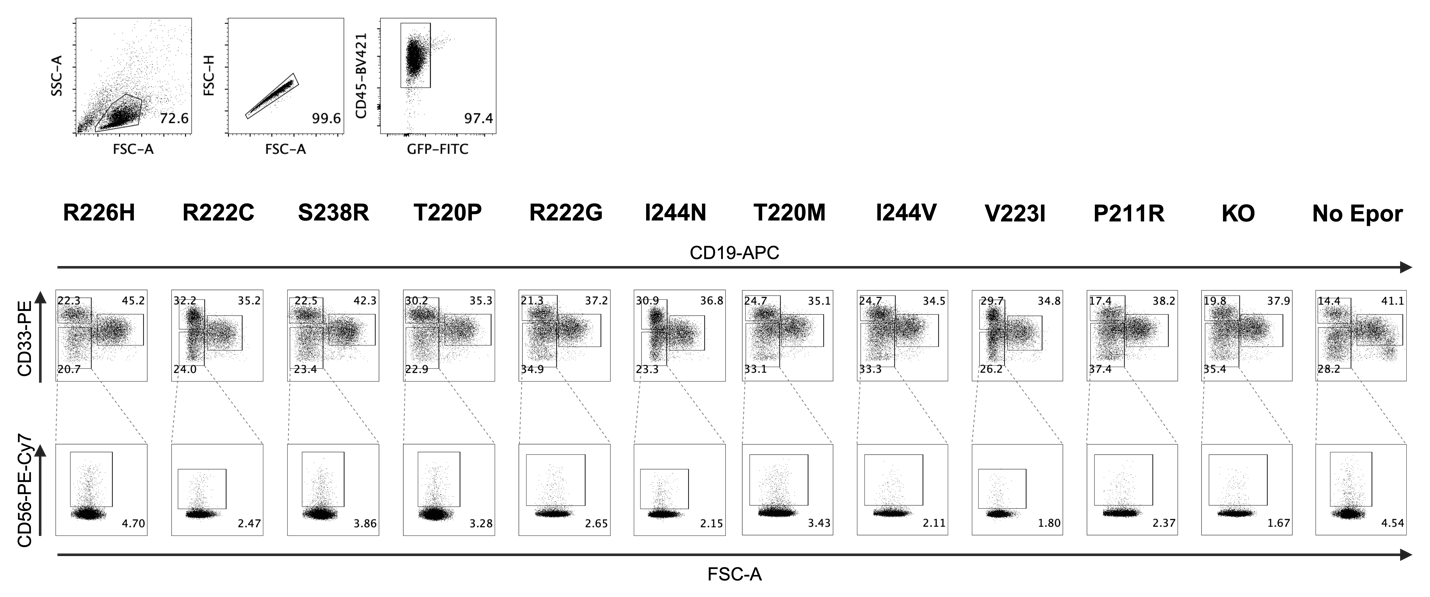

Figure S11: *Representative flow cytometry gating strategy and sort gates from Week 3 T-cell differentiation (top) and Week 3 B-cell differentiation (bottom) during HSPC validation studies of selected variants on interest or controls (KO = knockout, No Epor = treated similarly (“mock”) but without electroporation.*

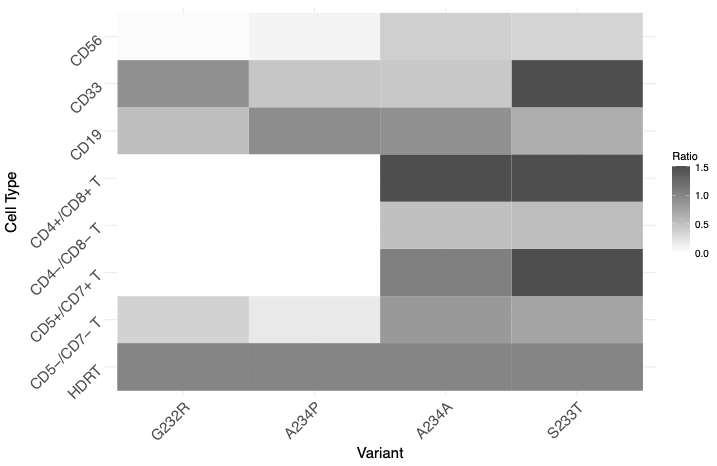

Figure S12

Figures S10-12: Two known or predicted pathogenic variants (G232R and A234P) and benign variants (A234A and S233T) were recreated in CD34+ cord blood HSPCs and validated as in Fig 2B by differentiation on the OP9-7FS coculture system into CD5+ CD7+ pro T cells at week 3, CD4+ CD8+ pre T cells at week 6, or on the OP9-R coculture system into CD19+ pre-B cell, CD33+ myeloid lineage, or CD56+ NK-cell progenitor populations with gating strategy as in SFig 7). Cells were sorted for surface markers of maturation along each respective lineage, then gDNA was extracted and sequenced to calculate the relative frequency of either variant allele. The functional impact of a given variant upon HSPC differentiation into respective lineages is measured by the prevalence of alleles bearing the variant of interest relative to the benign variant. Heatmap of adjusted ratios of alleles for selected test variants (relative to ClinVar benign variant (H236H), normalized to initial ratio in the HDR template) in sorted cell populations (SFig 11).

**Supplementary Discussion**

As X-SCID is defined by a failure of development of T and NK-cells, we expected to see an effect of blocked T-cell differentiation from HSPCs. Initially, we attempted to screen the full variant library directly in HSPCs differentiated *in vitro* using the OP9-DL1 co-culture system to generate CD4+CD8+ (DP) T cell progenitors (Supplementary Figure 7) ^46^. We found that although the nonsense mutations trended negative in comparison to synonymous (Wilcoxon test p=2.7x10^-5), random variant dropout and data sparsity most likely due to low library coverage in HSPCs - presumably due low HDR rates and lack of true T cell progenitors, resulting in edited HSPC clones that never fully differentiate - created considerable signal noise, causing synonymous mutations to appear damaging (Supplementary Figure 7,8,9). We thus opted for a focused arrayed validation of novel screen-identified variants similar to the validation studies in T cells.

Our follow-up experiments on individual variants in edited HSPCs revealed new insights that can be used for hypothesis generation. The variant I224V c.730A>G also showed a potential gain-of-function effect in the CD5+/7+ and NK cell populations, which warrants further investigation (Figure 2D, S5). Interestingly, R222C c.664C>T, an annotated X-SCID mutation that has been associated with atypical “leaky” SCID phenotype (Tlow/− B+ NK+)^13^, was largely depleted in the CD5+/7+ cell population, comparable to annotated pathogenic and screen-predicted damaging variants (e.g., R226H) (Figure 2D).
